# Effects of AI-driven Lifestyle Intervention on Psychological Well-Being and Body Image Among Young Adults In Malaysia

**DOI:** 10.64898/2026.07.20.26358442

**Authors:** Amirah Amri, Izzul Azmi, Aidel Zafran, Nurin Adibah, Haikal Zulkafli, Alif Iman, Adam Linoby

## Abstract

**Background:** University students experience substantial psychological well-being and body-image concerns, while scalable, personalized digital support remains underexamined in Malaysia. Artificial intelligence chatbots may deliver repeated lifestyle guidance, but the incremental value of personalization over structured chatbot support is uncertain.

**Objectives:** This study evaluated changes in psychological well-being and body appreciation following a 12-week personalized AI-powered lifestyle intervention, NExGEN, among Malaysian university students.

**Methods:** A two-arm, controlled, quasi-experimental pre-post study allocated 140 students aged 18–35 years by matched blocks to NExGEN (*n* = 70) or a structured-prompt ChatGPT control (*n* = 70). NExGEN generated adaptive weekly lifestyle actions from a 47-item onboarding assessment, whereas control participants received standardized weekly prompts covering the same lifestyle domains. Psychological well-being and body appreciation were assessed at baseline and week 12 using the World Health Organization-Five Well-Being Index and Body Appreciation Scale-2. Intention-to-treat linear mixed models estimated adjusted within-group changes and between-group differences in change, with Holm adjustment for the co-primary outcomes.

**Results:** Week-12 assessments were completed by 121 participants (86.43%). In NExGEN, psychological well-being improved by an adjusted 8.68 points (95% CI, 6.22–11.14), *z* = 6.91, *p* < .001, and body appreciation improved by 0.17 points (95% CI, 0.10–0.24), *z* = 4.82, *p* < .001. However, between-group differences in change were not statistically significant for psychological well-being (2.87 points; 95% CI, −0.48 to 6.23; *z* = 1.68; Holm-adjusted *p* = .093) or body appreciation (0.10 points; 95% CI, 0.00–0.19; *z* = 1.99; Holm-adjusted *p* = .093). Median platform logins were 68.00 in NExGEN and 58.50 in control; mean acceptability scores were 3.92 and 3.59, respectively.

**Conclusions:** NExGEN participation was associated with significant within-group improvements in psychological well-being and body appreciation, but personalized guidance did not demonstrate superiority over structured chatbot guidance. Because allocation was quasi-experimental, causal attribution remains limited. Randomized component-level trials are needed to determine whether personalization provides incremental benefit.

**Highlights:**

- NExGEN improved WHO-5 scores by 8.68 points over 12 weeks.
- NExGEN improved BAS-2 scores by 0.17 points over 12 weeks.
- Between-group WHO-5 change was not significant (Holm p = .093).
- Between-group BAS-2 change was not significant (Holm p = .093).
- NExGEN acceptability averaged 3.92 of 5, versus 3.59 in control.

## 1. Introduction

University life coincides with a developmental period in which academic demands, financial concerns, changing social roles, and greater personal independence can place considerable pressure on psychological functioning (Nyakhar & Wang, 2025). In a multinational survey of 13,984 first-year university students, 31% met criteria for at least one common mental disorder during the previous 12 months, indicating that substantial mental-health needs are already present at entry into higher education (Auerbach et al., 2018). Malaysian evidence also signals a serious burden. During the COVID-19 period, a study of university students in Selangor reported moderate-to-severe symptoms of depression, anxiety, and stress in 53.9%, 66.2%, and 44.6% of respondents, respectively (Wong et al., 2023). These symptom estimates capture only one dimension of student well-being. Body image is also relevant because dissatisfaction with appearance may coexist with emotional difficulties and health-related behaviors (Hashim et al., 2022; Mahon & Seekis, 2022). Among Malaysian university students, longer social-networking use was associated with greater body dissatisfaction, with additional differences observed by sex and body mass index (Hashim et al., 2022). Together, these findings position psychological well-being and body image as connected concerns within the university population.

University mental-health support commonly relies on counselling, referral, and professionally delivered programs, yet the scale of student need can exceed the reach of services that depend on scheduled appointments and sustained face-to-face engagement. Cross-national evidence indicates that campus demand often exceeds available resources (Auerbach et al., 2018), while stigma, treatment costs, and limited access to providers can further discourage care (Nyakhar & Wang, 2025). These constraints have encouraged interest in approaches that can be accessed privately, repeatedly, and outside conventional service hours. For young adults accustomed to mobile communication, digitally delivered support may reduce practical barriers and provide timely guidance within everyday routines (Madrid-Cagigal et al., 2025). Nevertheless, accessibility alone does not ensure meaningful engagement or improvement. Interventions must also respond to individual needs, maintain relevance over time, and address outcomes that matter to students beyond clinical symptom reduction (Nyakhar & Wang, 2025).

Digital mental-health interventions have therefore become an important component of university well-being strategies. A recent systematic review and meta-analysis of 34 studies, including 21 randomized controlled trials, found medium pooled effects for depression and anxiety among university students, with fully automated interventions producing a larger pooled anxiety effect than guided formats (Madrid-Cagigal et al., 2025). Conversational agents extend this model by providing interactive responses rather than static educational content, allowing guidance to be delivered through dialogue across repeated exchanges (Abd-Alrazaq et al., 2020). Meta-analytic evidence indicates that artificial intelligence-based chatbots can produce small short-term reductions in depressive and anxiety symptoms, although effects may weaken after treatment and vary according to intervention duration and design (Zhong et al., 2024). Earlier evidence also suggested potential benefits of chatbots for selected mental-health outcomes, while emphasizing that findings for broader psychological well-being and safety remained limited (Abd-Alrazaq et al., 2020). These results support the promise of chatbot-delivered support but also show that technological availability should not be equated with established effectiveness across all dimensions of well-being.

The limitations become more apparent when the outcome focus of existing research is considered. A rapid systematic review of artificial intelligence chatbots for college students identified nine studies involving 1,082 participants; most reported improvements in at least one mental-health outcome, but heterogeneity, reliance on self-report, attrition reaching 61%, and limited generalizability weakened the certainty of conclusions (Nyakhar & Wang, 2025). Moreover, the literature has concentrated largely on depression, anxiety, stress, or distress, leaving positive psychological well-being and self-perception less consistently evaluated. Digital body-image interventions have developed as a separate field. A systematic review found that only eight of 15 interventions improved at least one body-image outcome, generally with small-to-medium effects and limited evidence of sustained change (Mahon & Seekis, 2022). A meta-analysis of 19 studies involving 2,424 adult women found small-to-moderate improvements in global body satisfaction and cognitive body dissatisfaction, but the evidence remained population-specific and was not centered on personalized chatbot-delivered lifestyle coaching (Conboy et al., 2024). Thus, evidence for digital mental-health support cannot automatically be extended to body image or integrated lifestyle outcomes.

Contextual transferability presents a further concern. Reviews of artificial intelligence chatbots have highlighted variation in intervention content, duration, comparison conditions, and participant characteristics, together with limited diversity across study populations (Nyakhar & Wang, 2025; Zhong et al., 2024). Consequently, evidence developed in other populations cannot be assumed to apply unchanged to Malaysian students, particularly for outcomes linked to self-perception and lifestyle. Existing Malaysian studies have primarily described psychological distress or body dissatisfaction rather than testing personalized digital interventions. The Selangor study, for example, identified high symptom levels and associations with exercise and other factors but did not evaluate an artificial intelligence-supported program (Wong et al., 2023). Similarly, Malaysian body-image evidence has linked social-networking exposure, sex, and body mass index with dissatisfaction without determining whether tailored lifestyle guidance can improve body-image perceptions over time (Hashim et al., 2022). Taken together, the available literature leaves two connected gaps: insufficient empirical evidence on personalized chatbot-delivered lifestyle modification for psychological well-being among Malaysian university students, and little evidence on whether such coaching can improve body image within the Malaysian sociocultural context.

Addressing these gaps requires evaluation of outcomes that extend beyond disorder symptoms and include both positive psychological functioning and body-image perception (Mahon & Seekis, 2022; Nyakhar & Wang, 2025). Accordingly, this quasi-experimental pre-post study aimed to determine the effects of a 12-week personalized AI-powered chatbot intervention, NExGEN, among young adult university students in Malaysia. The first objective was to evaluate changes in psychological well-being following the intervention, and the second was to evaluate changes in body image after 12 weeks. The study examined the extent to which NExGEN improved psychological well-being and influenced body-image perception from baseline to post-intervention. It was hypothesized that participants would demonstrate statistically significant improvements in psychological well-being and body image after completing the 12-week intervention compared with their baseline levels.

## 2. Methodology

### 2.1 1. Study Design and Setting

This study used a two-arm, controlled, quasi-experimental pre-post design with an embedded qualitative process evaluation. The quantitative component evaluated changes in psychological well-being and body image following a 12-week artificial intelligence-driven lifestyle intervention. Participants were assigned in a 1:1 ratio to either the NExGEN personalized intervention group or a structured-prompt control group. Outcome measurements were conducted at baseline and immediately after completion of the 12-week intervention. The qualitative component examined participants’ experiences of intervention personalization, engagement, acceptability, perceived benefits, unintended effects, and barriers to continued use.

Recruitment and data collection for the main study were conducted from April to June 2026. Baseline assessments and group allocation were completed on a rolling basis during this period. Each participant began the allocated intervention within seven days of completing the baseline assessment and followed an individual 12-week intervention schedule. Post-intervention assessments were conducted within seven days of each participant’s final intervention week.

Recruitment and physical assessments were conducted at three participating university campuses in the Seremban district of Negeri Sembilan, Malaysia. Assessments were performed in private university research rooms that provided sufficient space for questionnaire completion and anthropometric measurements. The NExGEN and control interventions were delivered remotely through participants’ internet-enabled smartphones, tablets, or computers. Baseline and week-12 assessment sessions lasted approximately 45 to 60 minutes.

Because the investigation was quasi-experimental, participants were not assigned using unrestricted individual randomization. Instead, matched block allocation was used to maintain comparability between groups during rolling recruitment. Allocation blocks consisted of four to six participants who completed baseline assessment during the same recruitment period. Participants were categorized according to sex, participating university, and baseline psychological well-being category. An independent statistician who was not involved in participant recruitment, intervention delivery, or outcome assessment prepared the allocation schedule. Participants were assigned sequentially to the group that maintained the closest balance across the prespecified matching variables. When two allocation options produced equivalent balance, a computer-generated random number was used to determine the assignment.

Recruitment staff submitted only the participant identification code and matching variables to the independent statistician after eligibility and baseline assessment had been completed. Group assignment was subsequently returned to the intervention coordinator through a password-protected file. Participants remained in their initially assigned groups throughout the study, regardless of intervention adherence or discontinuation.

Blinding of participants was not feasible because the NExGEN group received personalized and adaptive prompts, whereas the control group received a standardized library of general prompts. The intervention coordinator was also not blinded because this researcher provided the relevant platform instructions and monitored technical engagement. However, research personnel responsible for administering questionnaires and conducting physical measurements remained blinded to group allocation. Participants were instructed not to discuss the content, format, or personalization of their assigned chatbot intervention with outcome assessors.

Study groups were coded as Group A and Group B in the statistical dataset. The statistician received only the coded dataset and remained blinded to the meaning of the group codes until the primary analyses, assumption checks, and sensitivity analyses had been completed. Qualitative interviewers were not blinded because the interview guide required participants to describe their assigned intervention. Reporting was structured according to the Transparent Reporting of Evaluations with Nonrandomized Designs statement, which was developed for nonrandomized evaluations of behavioral and public-health interventions (Des Jarlais et al., 2004).

### 2.2 2. Participants and Recruitment

University students aged 18 to 35 years were eligible to participate. Participants were required to be enrolled in a diploma, undergraduate, or postgraduate program at one of the participating universities and to reside in Seremban or the surrounding commuting area throughout the 12-week study period. They were required to understand written Bahasa Melayu or English, possess an internet-enabled device, and have regular access to email or WhatsApp for study communication. Participants were also required to demonstrate sufficient digital literacy to open an internet link, enter information into a chatbot interface, and retrieve a previous conversation.

Students were eligible irrespective of sex, ethnicity, academic faculty, year of study, body mass index, or previous exposure to generative artificial intelligence. However, students were excluded if they reported an acute psychiatric crisis, active self-harm intent, or another condition requiring immediate clinical intervention. Students receiving stable psychological counseling or psychiatric treatment remained eligible when no major treatment change was anticipated during the intervention. Those who had initiated or substantially changed psychotropic medication during the preceding four weeks were excluded.

Students were also excluded if they reported a clinically managed eating disorder or another condition for which unsupervised lifestyle or body-image guidance could be inappropriate. Participation in another structured digital mental-health, body-image, weight-management, or lifestyle intervention was prohibited. Individuals who had participated in the preliminary feasibility study were excluded from the main study. Only one participant from each household was enrolled to reduce the likelihood of intervention contamination.

Eligibility was initially assessed through a secure online screening form. The form collected information on age, student status, university, residential location, preferred language, digital access, concurrent interventions, and relevant safety considerations. Potentially eligible students were contacted by a trained research assistant for a brief telephone verification. Students who remained eligible were provided with an electronic participant-information sheet and were given at least 48 hours to consider participation before scheduling a baseline appointment.

The required sample size was estimated using G*Power version 3.1.9.7 (Faul et al., 2009). The calculation was based on two intervention groups, two measurement occasions, an assumed correlation of 0.50 between baseline and post-intervention measurements, 80% statistical power, and a two-sided familywise significance level of 0.05. A small-to-moderate group-by-time effect equivalent to a standardized between-group difference in change of approximately 0.40 was assumed. Psychological well-being and body appreciation were treated as co-primary outcomes, with multiplicity managed using Holm’s sequential procedure.

The initial calculation indicated that approximately 120 participants with analyzable repeated measurements were required. The target was increased to 140 participants to allow for approximately 14% attrition, withdrawal, or incomplete week-12 assessment. The final recruitment target was therefore 70 participants per group. The 12 participants involved in the feasibility study were not included in the main sample or statistical analysis.

Participants were recruited through official university email lists, learning-management systems, student-affairs announcements, and verified university social-media channels. Printed posters containing a QR code were displayed at student centers, residential colleges, libraries, cafeterias, sports facilities, and faculty noticeboards. Student representative councils and university health units distributed the standardized recruitment material but did not identify or directly approach individual students. Lecturers were permitted to display a recruitment slide at the end of class but were not involved in eligibility determination or enrollment.

Interested students accessed the online screening form through the QR code or recruitment link. Research assistants contacted eligible students within three working days. Written informed consent and final eligibility confirmation were completed before baseline measurements. Recruitment proceeded on a rolling basis to allow participants to begin the intervention shortly after baseline assessment.

Each participant received a unique study identification code. Enrollment records documented the recruitment source, eligibility outcome, reason for exclusion, consent status, baseline attendance, group assignment, intervention initiation, and assessment completion. Participants received reimbursement for verified local travel to the baseline and week-12 assessments. No performance-based or outcome-based incentives were provided. Access to the digital intervention platform was provided without charge throughout the study period.

### 2.3 3. Intervention Protocols

#### NExGEN Intervention

Participants assigned to the intervention group received the NExGEN AI-powered personalized lifestyle intervention for 12 weeks. NExGEN was configured as a secure web-based prompt-generation platform, version 2.1, connected to ChatGPT model 5.4. The generative model was accessed through a study-managed institutional account rather than participants’ personal accounts. External web browsing, third-party plug-ins, file uploads, and image-generation functions were disabled.

The NExGEN system used a fixed researcher-developed system instruction and standardized response parameters. The model temperature was set at 0.4 to limit unnecessary response variability, while maximum response length was restricted to maintain concise and manageable guidance. Participants selected either Bahasa Melayu or English during onboarding.

Personalization was based on a 47-item onboarding questionnaire covering demographic characteristics, academic schedule, usual sleep pattern, physical activity, dietary routine, perceived stressors, body-image concerns, available social support, environmental resources, personal barriers, previous lifestyle-change attempts, and preferred coaching style. Participants were instructed not to enter their names, student numbers, addresses, medical-record information, or identifiable information concerning other individuals.

NExGEN converted the onboarding responses into a structured personal-context prompt and generated an individualized 12-week lifestyle-coaching framework. The framework addressed psychological well-being, stress self-management, sleep regularity, safe physical activity, meal organization, social connectedness, self-compassion, and positive body-image practices. The system was explicitly instructed not to diagnose psychiatric or medical conditions, recommend medication, provide psychotherapy, or prescribe extreme dietary restriction.

At the beginning of each intervention week, NExGEN generated an individualized plan containing two to four achievable actions. The recommended actions were adjusted to the participant’s academic timetable, perceived barriers, available resources, previous adherence, and stated preferences. Participants completed a brief digital check-in at the end of each week, reporting progress, perceived difficulty, mood, barriers, and preferred adjustments. These responses were incorporated into the subsequent week’s personalized prompts.

Participants were asked to interact with ChatGPT on at least three days per week. A typical interaction lasted approximately 10 to 15 minutes. Automated reminders were issued after three consecutive days without a recorded interaction. Research staff provided technical assistance but did not provide additional lifestyle, psychological, dietary, or exercise coaching. All technical assistance followed a standardized script.

Engagement information was obtained from platform logs and included login dates, number of interactions, prompt categories, weekly check-in completion, response length, and intervention discontinuation. Intervention fidelity was reviewed weekly using automated reports and a randomly selected 10% sample of de-identified chatbot exchanges. Any platform update or prompt modification was documented through version control before implementation.

#### Control Condition

Participants assigned to the control group received a structured manual-prompt program delivered through the same ChatGPT model and institutional access environment. The control condition covered the same broad lifestyle domains and was delivered over the same 12-week period. Participants received standardized prompts related to stress management, sleep hygiene, physical activity, balanced eating, study-life organization, general self-care, and body respect.

The control prompts did not use participants’ onboarding responses and did not construct an individualized longitudinal profile. The wording, sequence, and timing of the core prompts were identical for all control participants. Participants received one prompt package each week and were instructed to interact with the chatbot on at least three days per week. They could ask clarification questions, but the system instruction prevented ChatGPT from adapting future weekly content to their personal progress or barriers.

Control participants completed brief weekly check-ins to document prompt use and technical difficulties. Their responses did not alter the subsequent prompt package. The control group received the same orientation duration, platform-access period, reminder frequency, technical-support procedures, and safety-monitoring procedures as the NExGEN group. No human lifestyle counseling was routinely provided.

#### Standardization and Intervention Fidelity

Both groups received the same broad healthy-lifestyle objectives so that the principal difference between conditions was the level of personalization. Participants were encouraged to establish a regular sleep schedule, increase safe physical activity where appropriate, maintain consistent meals, consume adequate fluids, undertake brief stress-management activities, and reduce appearance-based social comparison. Body-image guidance emphasized body functionality, respect, self-compassion, and avoidance of stigmatizing weight-related language.

No mandatory weight-loss target was imposed because psychological well-being and body image were the principal study outcomes. Participants could independently select weight-related goals, but the intervention systems were instructed not to recommend rapid weight loss, punitive exercise, severe energy restriction, or unsafe supplementation.

A standardized 30-minute orientation was provided during the baseline visit. The orientation included a live platform demonstration, a written user guide, a brief instructional video, safety information, and a practice interaction. Intervention exposure was defined as completion of at least 70% of the expected weekly check-ins and use of the assigned chatbot on at least two days during eight of the 12 intervention weeks.

Participants who did not meet the adherence threshold remained included in the intention-to-treat analysis but were excluded from the per-protocol sensitivity analysis. Ordinary university wellness services, medical treatment, and existing counseling were permitted. Initiation of another structured digital lifestyle intervention was discouraged and documented as a potential protocol deviation.

Potentially harmful chatbot content was monitored through automated screening for language associated with self-harm, severe psychological distress, eating-disorder risk, and unsafe health practices. Flagged interactions generated a standardized safety message and were reviewed by the designated safety officer within one working day. Statements indicating immediate risk triggered direct contact and implementation of the ethics-approved emergency referral protocol.

### 2.4 4. Outcome Variables

Psychological well-being and body appreciation were the two co-primary outcomes. Both outcomes were assessed at baseline before group allocation and at week 12 after intervention completion. The primary psychological well-being estimand was the adjusted between-group difference in change in well-being score from baseline to week 12. The primary body-image estimand was the adjusted between-group difference in change in body-appreciation score over the same period.

#### Psychological Well-Being

Psychological well-being was assessed using the World Health Organization-Five Well-Being Index. The WHO-5 consisted of five positively worded items assessing subjective well-being during the preceding two weeks. Each item was scored from 0, indicating “at no time,” to 5, indicating “all of the time.” Item scores were summed to produce a raw score from 0 to 25 and multiplied by four to generate a standardized score from 0 to 100. Higher scores represented better psychological well-being (Topp et al., 2015).

Participants selected either the English or validated Malay version during baseline assessment and completed the same language version at week 12. The Malay WHO-5 had undergone translation and psychometric evaluation in a Malaysian sample (Suhaimi et al., 2022).

#### Body Image

Body image was operationalized as positive body appreciation and measured using the Body Appreciation Scale-2. The BAS-2 consisted of 10 items rated on a five-point scale from 1, indicating “never,” to 5, indicating “always.” The total score was calculated as the mean of the completed items, producing a possible score from 1 to 5. Higher scores represented greater acceptance of, respect for, and appreciation of the body (Tylka & Wood-Barcalow, 2015).

Participants completed either the English or Bahasa Malaysia version. The Bahasa Malaysia translation had demonstrated a unidimensional structure and acceptable psychometric properties in Malaysian adults (Swami et al., 2019). The English version had also demonstrated evidence of validity and measurement invariance in a Malaysian sample (Tan et al., 2021).

#### Secondary and Process Outcomes

Secondary and process outcomes included intervention adherence, digital engagement, participant satisfaction, perceived usefulness, technical difficulties, and reported adverse experiences. Digital engagement was obtained directly from timestamped platform records. Participant satisfaction and perceived usefulness were assessed at week 12 using an eight-item study-specific questionnaire scored from 1 to 5.

Body weight was assessed as an exploratory physiological outcome using a calibrated seca mBCA 515 body-composition analyzer. Participants removed footwear, heavy outer clothing, belts, and pocket contents before measurement. They were instructed to avoid vigorous physical activity and alcohol for 24 hours, avoid food and caloric drinks for at least eight hours, and empty their bladder before assessment. Measurements were scheduled at a similar time of day at baseline and week 12 where feasible.

Two body-weight readings were obtained and recorded to the nearest 0.1 kg. A third reading was obtained when the first two differed by more than 0.2 kg, and the mean of the two closest readings was recorded. Height was measured once at baseline using a wall-mounted stadiometer and recorded to the nearest 0.1 cm. Body mass index was calculated as weight in kilograms divided by height in meters squared.

#### Assessment Procedures

Questionnaires were completed electronically in a private assessment room using password-protected tablets. Research assistants explained the response procedure but did not interpret questionnaire items for participants. The same questionnaire order and language version were maintained at baseline and week 12.

A WHO-5 or BAS-2 score was calculated when at least 80% of the relevant scale items had been completed. When one WHO-5 item or no more than two BAS-2 items were missing, the participant’s mean response across the completed items was substituted for the missing item or items. Scales with greater missingness were coded as missing and addressed through the prespecified missing-data procedure. Internal consistency was evaluated at each assessment using Cronbach’s alpha and McDonald’s omega.

### 2.5 5. Data Analysis

Quantitative data were managed and analyzed using IBM SPSS Statistics, Version 30. GraphPad Prism, Version 10.5, was used to prepare figures and visualize adjusted outcome estimates. G*Power, Version 3.1.9.7, was used for the sample-size calculation. Qualitative data were organized using NVivo 15.3.

Before analysis, the dataset was checked for duplicate records, impossible values, inconsistent coding, scoring errors, and values outside the prespecified range. All corrections were documented in a data-cleaning log. Statistical syntax, data dictionaries, imputation specifications, model outputs, and figure source files were retained in a version-controlled project directory. Analyses were conducted using a locked, de-identified dataset.

Continuous baseline variables were summarized using means and standard deviations when distributions were approximately symmetric and medians and interquartile ranges when distributions were markedly skewed. Categorical variables were summarized using frequencies and percentages. Baseline significance testing was not used to determine group comparability.

The primary analyses followed the intention-to-treat principle. Participants were analyzed in their allocated groups regardless of adherence, intervention discontinuation, or use of permitted concurrent services. Psychological well-being and body appreciation were analyzed separately using linear mixed models for repeated measures. Each model included fixed effects for group, time, and the group-by-time interaction. Participant identification was entered as a random intercept to account for the correlation between repeated measurements.

An unstructured covariance matrix was specified for the two assessment occasions. Models were adjusted for age, sex, participating university, previous use of ChatGPT, and the baseline value of the alternate co-primary outcome. The primary intervention effect was represented by the group-by-time interaction. Adjusted marginal means, within-group changes, between-group differences in change, standardized effect sizes, and 95% confidence intervals were reported.

Because assignment was quasi-experimental, a propensity score was estimated using age, sex, participating university, year of study, baseline WHO-5 score, baseline BAS-2 score, body mass index, previous chatbot use, and current counseling status. Stabilized inverse-probability-of-treatment weights were applied in a sensitivity analysis to evaluate the effect of measured allocation differences.

All tests were two-sided. The familywise significance level was set at 0.05. Holm’s sequential procedure was used to adjust the two co-primary outcome tests. Secondary and exploratory outcomes were interpreted primarily using effect estimates and confidence intervals.

Model assumptions were examined using residual-versus-fitted plots, quantile-quantile plots, influence statistics, and model-convergence information. Robust standard errors were calculated as a sensitivity analysis when residual distributions demonstrated substantial departure from model assumptions. Exploratory body-weight change was analyzed using the same group-by-time structure but was not included in the multiplicity-adjusted co-primary testing family.

A per-protocol sensitivity analysis included participants who completed the week-12 assessment, satisfied the prespecified intervention-exposure threshold, and had no major evidence of cross-group contamination. Engagement outcomes were summarized descriptively and explored as predictors of change within the NExGEN group. These engagement analyses were treated as exploratory.

#### Missing Data

The frequency, timing, pattern, and documented reasons for missing data were summarized by study group. Linear mixed models incorporated all available repeated measurements under a missing-at-random assumption. Multiple imputation by chained equations was conducted as the primary missing-data sensitivity analysis, with 50 imputed datasets generated. Multiple imputation was selected to account for uncertainty in unobserved values rather than replacing each missing value with a single fixed estimate (White et al., 2011).

The imputation model included group allocation, psychological well-being and body-appreciation scores at both measurement occasions, age, sex, university, year of study, body mass index, previous chatbot use, counseling status, intervention engagement, and recorded reasons for noncompletion. Continuous variables were imputed using predictive mean matching. Binary and categorical variables were imputed using logistic or multinomial logistic models, as appropriate.

The primary mixed models were fitted separately to each imputed dataset, and estimates were pooled using Rubin’s rules. Imputation diagnostics included trace plots, comparisons of observed and imputed distributions, and assessment of convergence. Complete-case analysis was performed as an additional sensitivity analysis. A pattern-mixture sensitivity analysis applied progressively less favorable week-12 values to participants with missing outcomes to examine departures from the missing-at-random assumption. Last observation carried forward was not used.

### 2.6 6. Ethical Considerations

Ethical approval was obtained from the Universiti Teknologi MARA Research Ethics Committee under reference REC/1954/2026. Administrative permission was obtained from each participating university before recruitment began. The approved protocol covered digital intervention delivery, questionnaire administration, physical measurement, chatbot-data handling, safety monitoring, qualitative interviews, and secondary use of de-identified data.

Potential participants received a detailed information sheet at least 48 hours before baseline assessment. The information sheet described the study objectives, group-allocation procedure, 12-week commitment, questionnaires, body measurements, digital monitoring, qualitative interview option, foreseeable risks, possible benefits, confidentiality arrangements, and withdrawal rights. Written informed consent was obtained before final eligibility confirmation and baseline data collection.

Participants were informed that participation was voluntary and that declining or withdrawing would not affect their academic grades, university services, healthcare, or relationship with university staff. Participants could discontinue chatbot use while allowing previously collected data to be retained and could still complete the week-12 assessment.

The consent process emphasized that participants should not enter names, student numbers, addresses, medical-record information, or identifiable information concerning third parties into the chatbot. Participants could request deletion of identifiable chatbot records before de-identification and dataset locking.

A unique study code was used on all study records. The file linking participant identities with study codes was stored separately from the research dataset on an encrypted institutional server. Access to identifiable information was restricted to the principal investigator and designated data manager.

Information transmitted between the NExGEN interface, institutional server, and language-model service was encrypted. Only the minimum information required for prompt personalization was transferred to the language-model service. Direct identifiers were not included in chatbot prompts. Chat transcripts were de-identified before fidelity or safety review and were not stored on personal computers, removable drives, or consumer cloud-storage services.

An independent safety officer with mental-health training reviewed reports of severe psychological distress, self-harm risk, suspected disordered eating, or unsafe chatbot recommendations. The chatbot displayed a persistent notice explaining that it did not replace medical, psychological, nutritional, or emergency care. Participants requiring additional support were referred to university counseling services, an appropriate healthcare provider, or emergency services according to the assessed level of risk.

De-identified quantitative data, analysis files, consent documentation, and audit records were retained for seven years after publication and were scheduled for secure destruction thereafter. Reports and publications presented only aggregated findings. The study received no funding.

#### Preliminary Feasibility Study

A separate preliminary feasibility study was conducted with 12 university students, including six assigned to NExGEN and six assigned to the structured-prompt control condition. The feasibility study was conducted before the main data-collection period, which ran from April to June 2026.

Pilot participants met the same core requirements for age, student status, residence, language, digital access, and safety as participants in the main study. The feasibility study examined recruitment procedures, screening completion, understanding of consent, onboarding duration, chatbot access, reminder delivery, questionnaire usability, safety-flag procedures, physical-measurement workflow, data export, and interview-guide clarity.

Feasibility criteria included successful platform access by at least 10 of the 12 participants, completion of at least three of the four weekly check-ins by most participants, completion of baseline and follow-up assessment by at least 10 participants, and absence of unresolved critical safety failures. Brief cognitive interviews were conducted to identify unclear questionnaire wording and difficulties using the digital platform.

Pilot chatbot records were reviewed for inappropriate personalization, repetitive outputs, culturally unsuitable guidance, unsafe recommendations, and accidental collection of direct identifiers. The pilot resulted in simplified onboarding instructions, removal of duplicated onboarding questions, clarification of the control prompt schedule, and addition of a permanently visible crisis-support message. No pilot participant was included in the main study, and pilot outcome data were not combined with the main dataset. The intervention version, assessment manual, safety workflow, data dictionary, and platform configuration were finalized before main-study recruitment.

## 3. Results

### 3.1 Participant Flow and Analysis Populations

#### Participant flow, attrition, protocol deviations, and missing outcome data

Of 188 individuals screened, 180 (95.74%) completed screening, of whom 153 (85.00%) were eligible; 27 were ineligible and 8 withdrew before an eligibility decision. Of the eligible individuals, 140 (91.50%) consented, completed baseline assessment, were allocated equally to NExGEN and control, and initiated their assigned intervention; 13 declined consent (Figure 1). Week-12 assessments were completed by 121 of 140 participants (86.43%), including 61 of 70 in NExGEN (87.14%) and 60 of 70 in control (85.71%).

**Figure 1.**
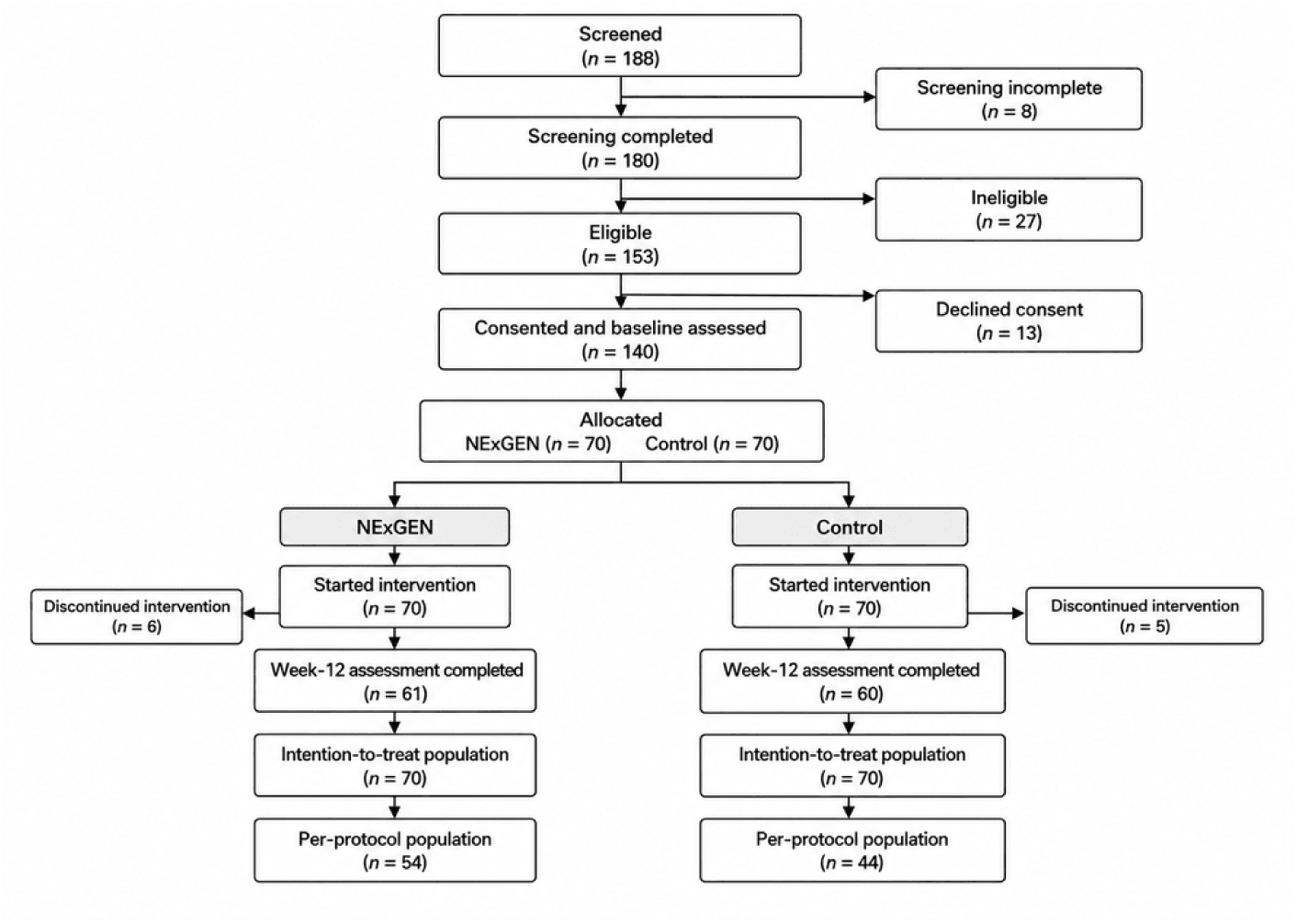
Participant flow through screening, allocation, intervention, follow-up, and analysis.

### 3.2 Participant Characteristics and Measurement Quality

#### A2 · Baseline demographic, academic, clinical, technological, and anthropometric characteristics

Baseline characteristics were available for all 140 participants, including 70 participants in the NExGEN group and 70 in the control group. The median age was 22.00 years (IQR, 21.00–24.00), while the mean height, body weight, and body mass index were 164.90 cm (SD, 8.53), 66.66 kg (SD, 14.40), and 24.36 kg/m² (SD, 3.99), respectively. Group-specific characteristics are presented in Table 1.

**Table 1.**
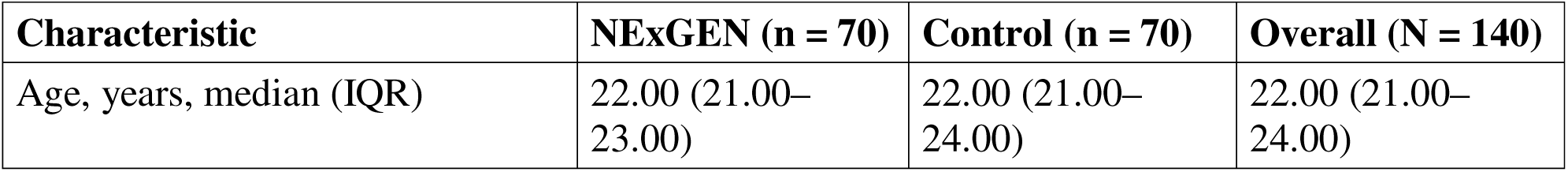
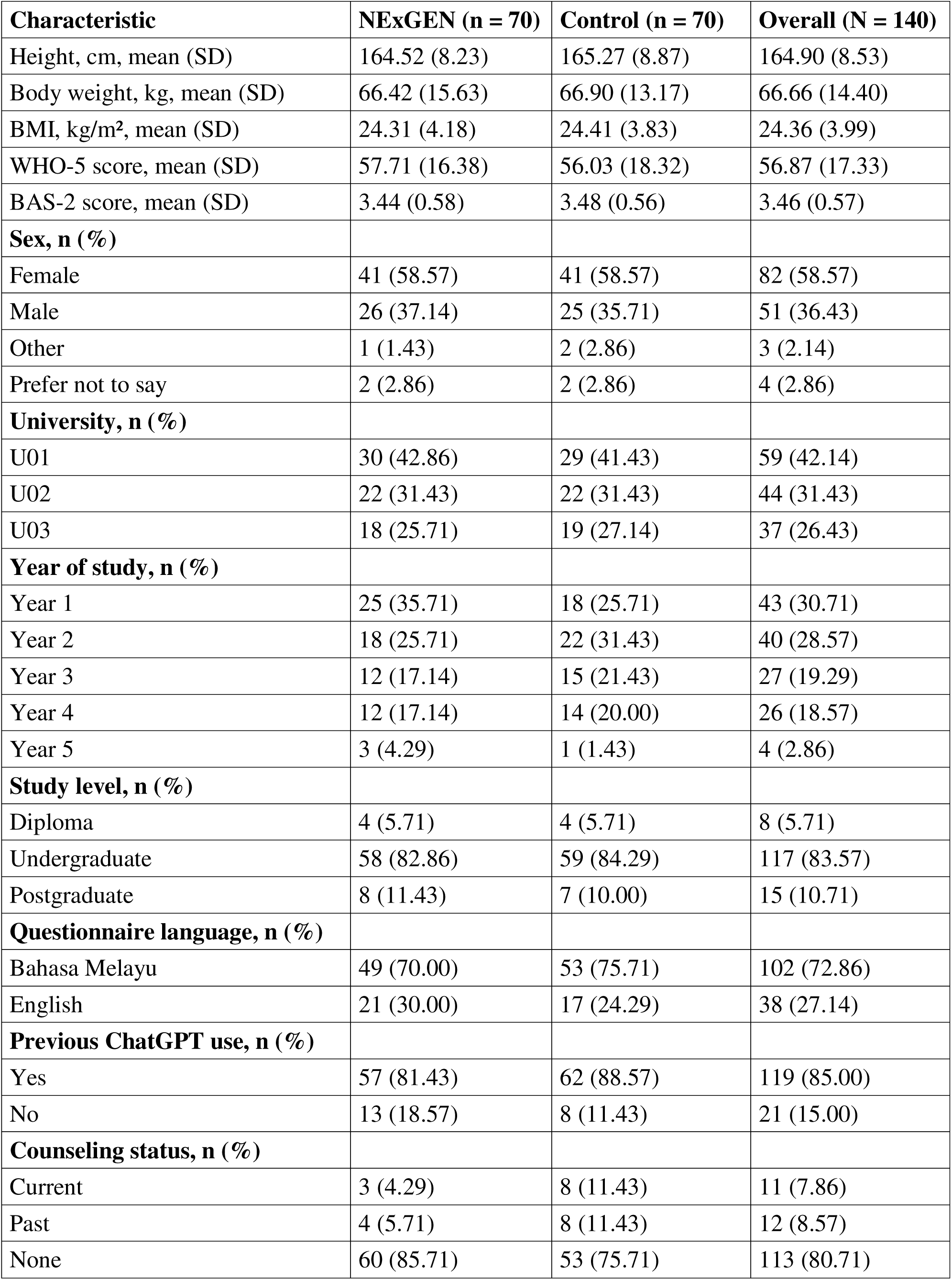
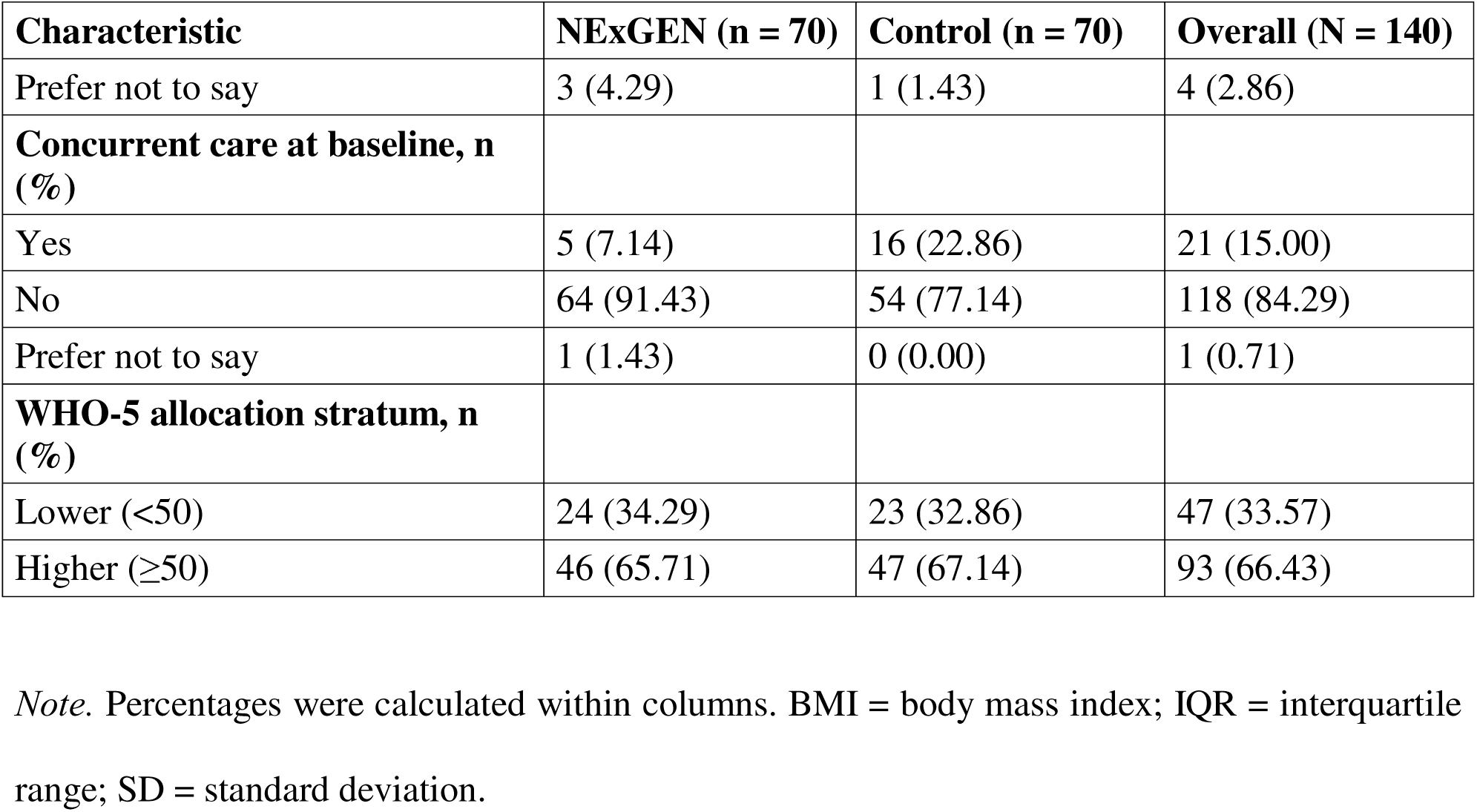
Baseline characteristics of participants by study group.

| Characteristic | NExGEN (n = 70) | Control (n = 70) | Overall (N = 140) |
| --- | --- | --- | --- |
| Age, years, median (IQR) | 22.00 (21.00–23.00) | 22.00 (21.00–24.00) | 22.00 (21.00–24.00) |

| **Characteristic** | **NExGEN (n = 70)** | **Control (n = 70)** | **Overall (N = 140)** |
| --- | --- | --- | --- |
| Height, cm, mean (SD) | 164.52 (8.23) | 165.27 (8.87) | 164.90 (8.53) |
| Body weight, kg, mean (SD) | 66.42 (15.63) | 66.90 (13.17) | 66.66 (14.40) |
| BMI, kg/m<sup>2</sup>, mean (SD) | 24.31 (4.18) | 24.41 (3.83) | 24.36 (3.99) |
| WHO-5 score, mean (SD) | 57.71 (16.38) | 56.03 (18.32) | 56.87 (17.33) |
| BAS-2 score, mean (SD) | 3.44 (0.58) | 3.48 (0.56) | 3.46 (0.57) |
| **Sex, n (%)** |  |  |  |
| Female | 41 (58.57) | 41 (58.57) | 82 (58.57) |
| Male | 26 (37.14) | 25 (35.71) | 51 (36.43) |
| Other | 1 (1.43) | 2 (2.86) | 3 (2.14) |
| Prefer not to say | 2 (2.86) | 2 (2.86) | 4 (2.86) |
| **University, n (%)** |  |  |  |
| U01 | 30 (42.86) | 29 (41.43) | 59 (42.14) |
| U02 | 22 (31.43) | 22 (31.43) | 44 (31.43) |
| U03 | 18 (25.71) | 19 (27.14) | 37 (26.43) |
| **Year of study, n (%)** |  |  |  |
| Year 1 | 25 (35.71) | 18 (25.71) | 43 (30.71) |
| Year 2 | 18 (25.71) | 22 (31.43) | 40 (28.57) |
| Year 3 | 12 (17.14) | 15 (21.43) | 27 (19.29) |
| Year 4 | 12 (17.14) | 14 (20.00) | 26 (18.57) |
| Year 5 | 3 (4.29) | 1 (1.43) | 4 (2.86) |
| **Study level, n (%)** |  |  |  |
| Diploma | 4 (5.71) | 4 (5.71) | 8 (5.71) |
| Undergraduate | 58 (82.86) | 59 (84.29) | 117 (83.57) |
| Postgraduate | 8 (11.43) | 7 (10.00) | 15 (10.71) |
| **Questionnaire language, n (%)** |  |  |  |
| Bahasa Melayu | 49 (70.00) | 53 (75.71) | 102 (72.86) |
| English | 21 (30.00) | 17 (24.29) | 38 (27.14) |
| **Previous ChatGPT use, n (%)** |  |  |  |
| Yes | 57 (81.43) | 62 (88.57) | 119 (85.00) |
| No | 13 (18.57) | 8 (11.43) | 21 (15.00) |
| **Counseling status, n (%)** |  |  |  |
| Current | 3 (4.29) | 8 (11.43) | 11 (7.86) |
| Past | 4 (5.71) | 8 (11.43) | 12 (8.57) |
| None | 60 (85.71) | 53 (75.71) | 113 (80.71) |
| Prefer not to say | 3 (4.29) | 1 (1.43) | 4 (2.86) |
| **Concurrent care at baseline, n (%)** |  |  |  |
| Yes | 5 (7.14) | 16 (22.86) | 21 (15.00) |
| No | 64 (91.43) | 54 (77.14) | 118 (84.29) |
| Prefer not to say | 1 (1.43) | 0 (0.00) | 1 (0.71) |
| **WHO-5 allocation stratum, n (%)** |  |  |  |
| Lower (<50) | 24 (34.29) | 23 (32.86) | 47 (33.57) |
| Higher (≥50) | 46 (65.71) | 47 (67.14) | 93 (66.43) |

*Note.* Percentages were calculated within columns. BMI = body mass index; IQR = interquartile range; SD = standard deviation.

#### Internal consistency of the WHO-5 and BAS-2

For the WHO-5, baseline reliability was α = 0.88 (95% bootstrap CI, 0.85–0.91) and ω = 0.89 (95% bootstrap CI, 0.85–0.91) among 138 participants. At week 12, WHO-5 reliability was α = 0.90 (95% bootstrap CI, 0.86–0.92) and ω = 0.90 (95% bootstrap CI, 0.87–0.93) among 114 participants. At week 12, BAS-2 reliability was α = 0.92 (95% bootstrap CI, 0.89–0.93) and ω = 0.92 (95% bootstrap CI, 0.89–0.93) among 111 participants.

### 3.3 Co-primary Outcomes

#### Psychological well-being: WHO-5 change from baseline to week 12

The intention-to-treat analysis included 258 WHO-5 observations from 140 participants. Baseline data were available for 70 NExGEN and 70 control participants, while week-12 data were available for 58 NExGEN and 60 control participants; 22 week-12 WHO-5 outcomes were unavailable and were not imputed. The model-estimated WHO-5 mean in the NExGEN group increased from 58.31 (95% CI, 55.25–61.37) at baseline to 66.99 (95% CI, 63.22–70.75) at week 12, representing a change of 8.68 points (95% CI, 6.22–11.14), z = 6.91, p < .001. The model-estimated control-group mean increased from 55.43 (95% CI, 52.07–58.79) to 61.24 (95% CI, 57.83–64.65), representing a change of 5.80 points (95% CI, 3.53–8.08), z = 4.99, p < .001. Th difference in change between groups was 2.87 points (95% CI, −0.48 to 6.23), z = 1.68, raw p = .093 and Holm-corrected p = .093. The standardized difference in change, calculated using the pooled baseline standard deviation, was 0.17 (95% CI, −0.03 to 0.36). Leave-one-participant estimates of the group-by-time contrast ranged from 2.46 to 3.56 points (Figure 2).

**Figure 2.**
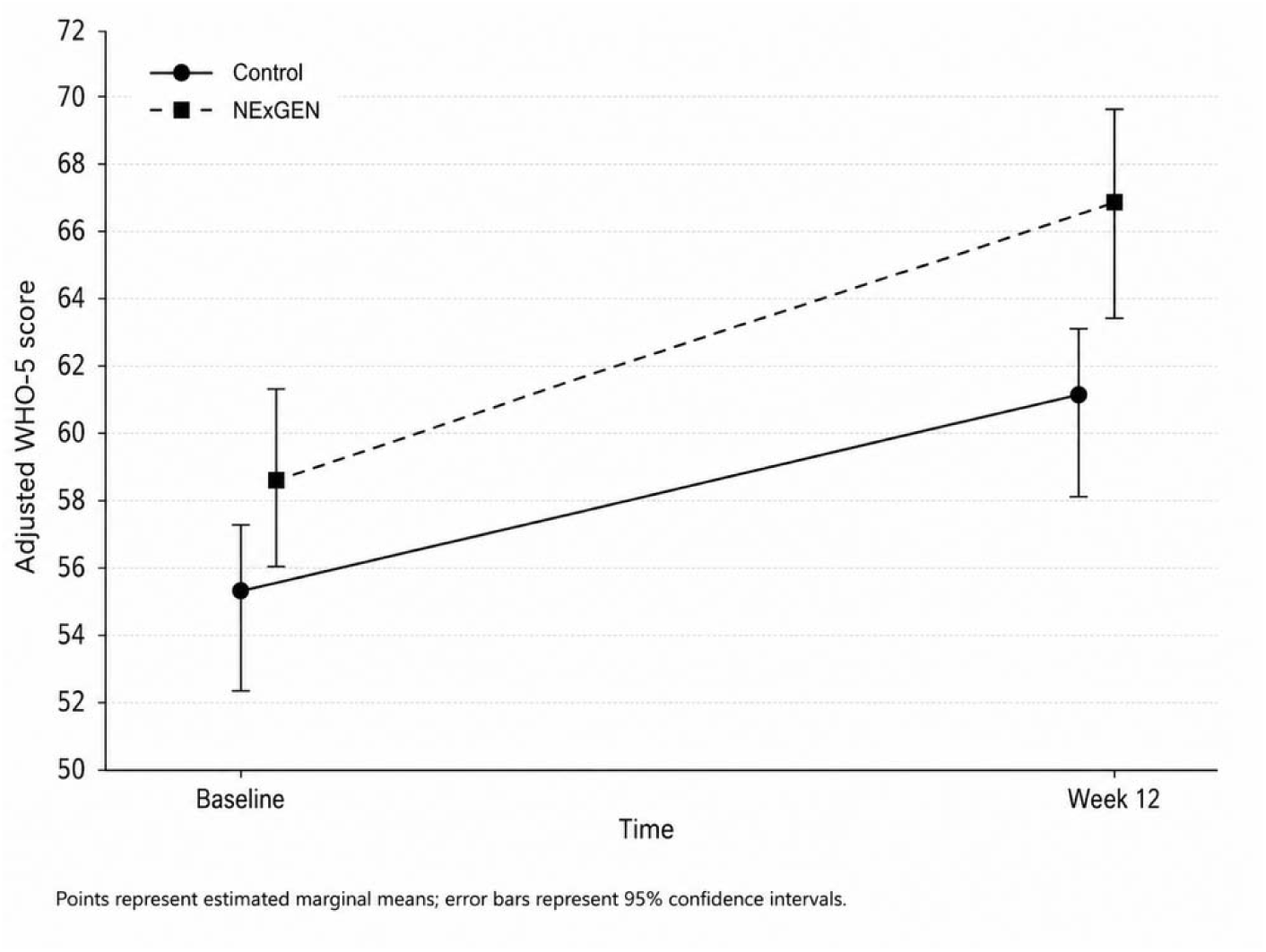
WHO-5 scores at baseline and week 12 by study group. Points represent estimated marginal means; error bars represent robust 95% confidence intervals.

#### Body image: BAS-2 change from baseline to week 12

The intention-to-treat analysis included 257 BAS-2 observations from 140 participants. Baseline data were available for 70 NExGEN and 70 control participants, while week-12 data were available for 58 NExGEN and 59 control participants; 23 week-12 BAS-2 outcomes were unavailable and were not imputed. The model-estimated BAS-2 mean in the NExGEN group increased from 3.42 (95% CI, 3.31–3.53) at baseline to 3.59 (95% CI, 3.47–3.72) at week 12, representing a change of 0.17 points (95% CI, 0.10–0.24), z = 4.82, p < .001. The model-estimated control-group mean increased from 3.50 (95% CI, 3.39–3.61) to 3.57 (95% CI, 3.45–3.70), representing a change of 0.07 points (95% CI, 0.01–0.14), z = 2.24, p = .025. The difference in change between groups was 0.10 points (95% CI, 0.00–0.19), z = 1.99, raw p = .047 and Holm-corrected p = .093. The standardized difference in change, calculated using the pooled baseline standard deviation, was 0.17 (95% CI, 0.00–0.33). Leave-one-participant estimates of the group-by-time contrast ranged from 0.09 to 0.11 points (Figure 3).

**Figure 3.**
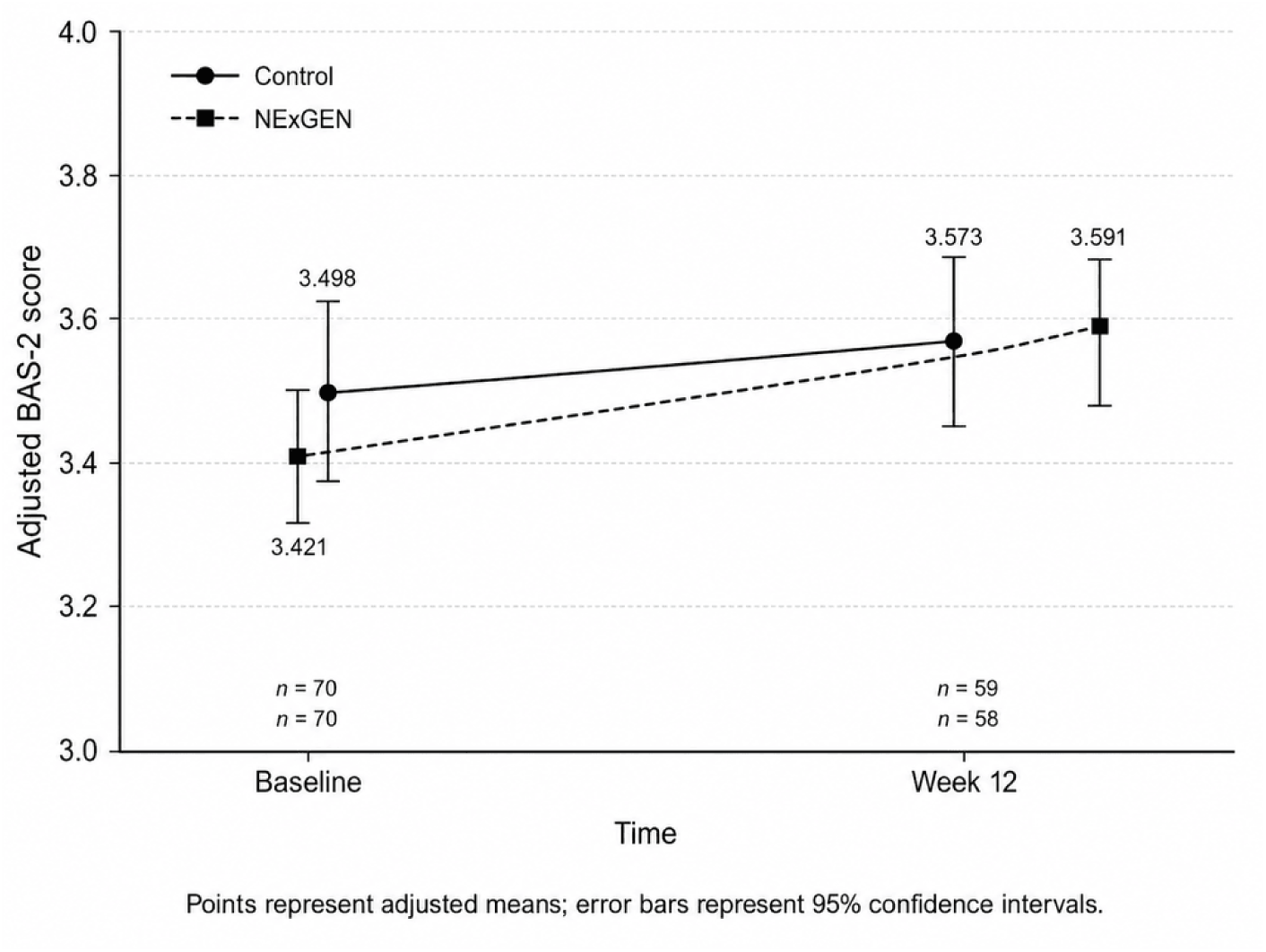
BAS-2 scores at baseline and week 12 by study group. Points represent estimated marginal means from models including age, sex, university, previous ChatGPT use, and baseline WHO-5 score; error bars represent robust 95% confidence intervals.

### 3.4 Intervention Delivery, Acceptability, and Safety

#### Intervention adherence and digital engagement

Participants in the NExGEN group recorded a median of 68.00 logins (IQR, 59.00–74.00), 38.00 active interaction days (IQR, 34.00–40.75), and 166.50 chatbot interactions (IQR, 150.00–198.00). Corresponding control-group values were 58.50 logins (IQR, 52.00–70.50), 34.00 active days (IQR, 29.00–38.00), and 141.00 interactions (IQR, 114.25–168.50). Prompt-category exposure by group is presented in Table 2.

**Table 2.**
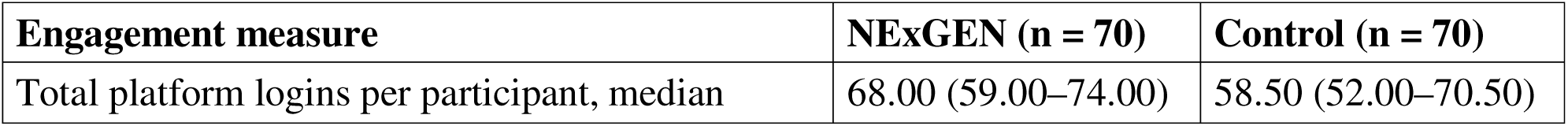
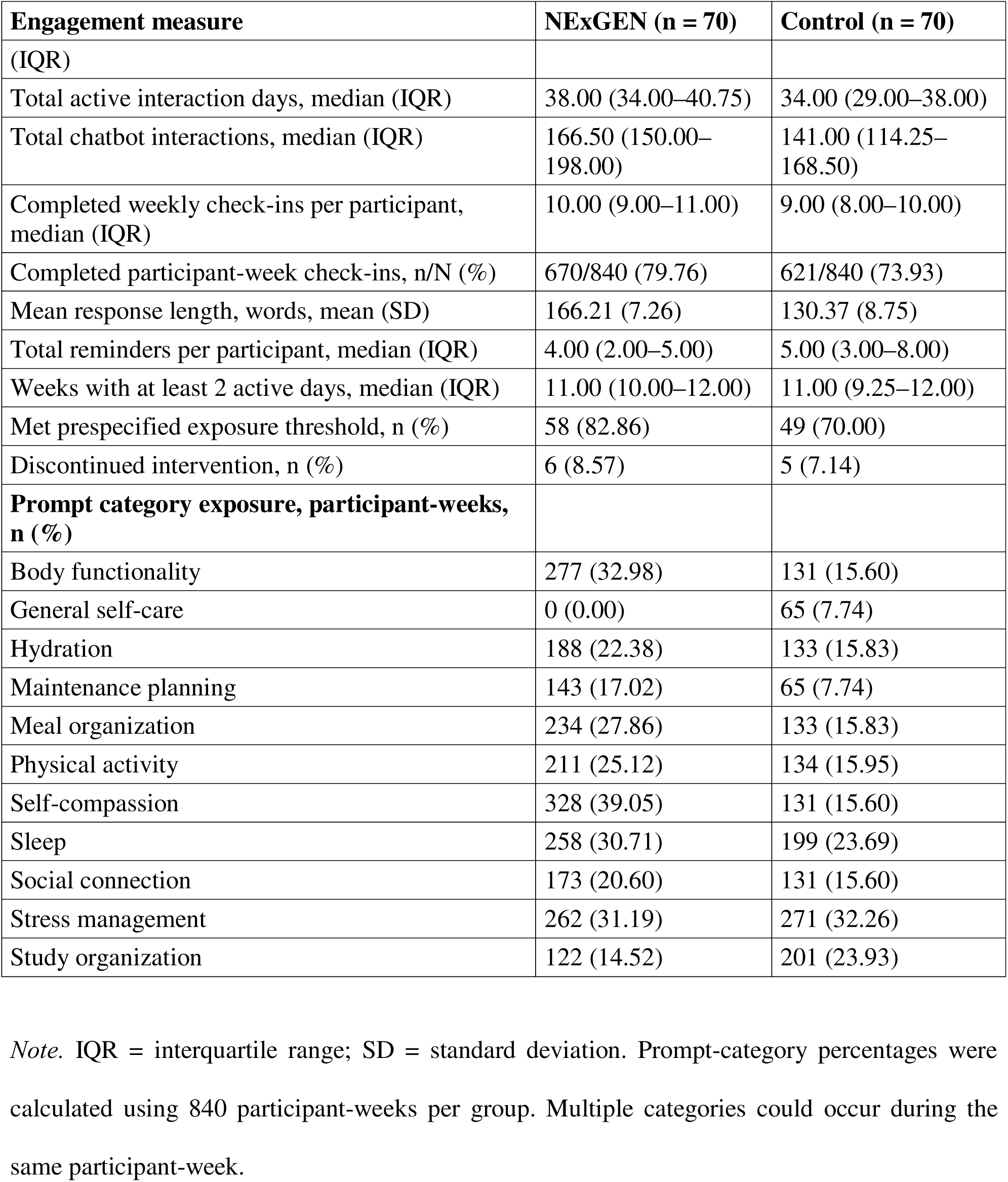
Intervention adherence and digital engagement by study group.

| Engagement measure | NExGEN (n = 70) | Control (n = 70) |
| --- | --- | --- |
| Total platform logins per participant, median | 68.00 (59.00–74.00) | 58.50 (52.00–70.50) |

| **Engagement measure** | **NExGEN (n = 70)** | **Control (n = 70)** |
| --- | --- | --- |
| (IQR) |  |  |
| Total active interaction days, median (IQR) | 38.00 (34.00–40.75) | 34.00 (29.00–38.00) |
| Total chatbot interactions, median (IQR) | 166.50 (150.00–198.00) | 141.00 (114.25–168.50) |
| Completed weekly check-ins per participant, median (IQR) | 10.00 (9.00–11.00) | 9.00 (8.00–10.00) |
| Completed participant-week check-ins, n/N (%) | 670/840 (79.76) | 621/840 (73.93) |
| Mean response length, words, mean (SD) | 166.21 (7.26) | 130.37 (8.75) |
| Total reminders per participant, median (IQR) | 4.00 (2.00–5.00) | 5.00 (3.00–8.00) |
| Weeks with at least 2 active days, median (IQR) | 11.00 (10.00–12.00) | 11.00 (9.25–12.00) |
| Met prespecified exposure threshold, n (%) | 58 (82.86) | 49 (70.00) |
| Discontinued intervention, n (%) | 6 (8.57) | 5 (7.14) |
| **Prompt category exposure, participant-weeks, n (%)** |  |  |
| Body functionality | 277 (32.98) | 131 (15.60) |
| General self-care | 0 (0.00) | 65 (7.74) |
| Hydration | 188 (22.38) | 133 (15.83) |
| Maintenance planning | 143 (17.02) | 65 (7.74) |
| Meal organization | 234 (27.86) | 133 (15.83) |
| Physical activity | 211 (25.12) | 134 (15.95) |
| Self-compassion | 328 (39.05) | 131 (15.60) |
| Sleep | 258 (30.71) | 199 (23.69) |
| Social connection | 173 (20.60) | 131 (15.60) |
| Stress management | 262 (31.19) | 271 (32.26) |
| Study organization | 122 (14.52) | 201 (23.93) |

*Note.* IQR = interquartile range; SD = standard deviation. Prompt-category percentages were calculated using 840 participant-weeks per group. Multiple categories could occur during the same participant-week.

#### Intervention fidelity and cross-group contamination

The fidelity log contained 187 records, comprising 24 automated reports, 159 sampled chatbot-exchange audits, and 4 version-control records. Six of the 159 sampled exchanges contained a deviation (3.77%): 4 of 74 NExGEN exchanges (5.41%) and 2 of 85 control exchanges (2.35%). All 6 detected deviations were recorded as resolved, with a median resolution time of 2.00 days (range, 2–2).

#### Participant satisfaction and perceived usefulness

Of 70 participants in each group, 61 NExGEN participants (87.14%) and 60 control participants (85.71%) provided scoreable acceptability questionnaires. The mean acceptability score was 3.92 (SD, 0.42) in NExGEN and 3.59 (SD, 0.41) in control. Complete responses to all questionnaire items were available for 118 participants; 3 participants had one missing item, and 19 did not complete the questionnaire.

#### Technical difficulties, adverse experiences, and concurrent support

Across 140 participants, 32 technical, safety, or concurrent-support events were recorded for 30 participants (21.43%). Thirteen events involved 13 of 70 NExGEN participants (18.57%), while 19 events involved 17 of 70 control participants (24.29%). Overall, 31 events (96.88%) were resolved and 1 distress-related event (3.13%) remained ongoing at database lock.

## 4. Discussion

Participation in the 12-week NExGEN intervention was associated with statistically significant within-group improvements in both co-primary outcomes. Psychological well-being increased by an adjusted 8.68 points (95% CI, 6.22–11.14), *z* = 6.91, *p* < .001, and body appreciation increased by an adjusted 0.17 points (95% CI, 0.10–0.24), *z* = 4.82, *p* < .001. These results supported the study hypotheses as prespecified within-group statements. They did not, however, establish that personalized NExGEN guidance was superior to structured, non-personalized chatbot guidance. The adjusted between-group difference in psychological well-being change was 2.87 points (95% CI, −0.48 to 6.23), *z* = 1.68, Holm-adjusted *p* = .093, while the corresponding difference in body appreciation change was 0.10 points (95% CI, 0.00–0.19), *z* = 1.99, Holm-adjusted *p* = .093. Both research-question contrasts therefore remained null after multiplicity adjustment. This pattern aligns with reviews showing that digital interventions can improve psychological outcomes among university students, while comparative advantages become less consistent when the comparator is itself an active intervention rather than no treatment or a wait-list condition (Davies et al., 2014; Harrer et al., 2019; Lattie et al., 2019). The study extends this literature by evaluating a personalized, generative-AI lifestyle intervention in Malaysian university students and by treating positive body appreciation as a co-primary outcome. It does not establish comparative effectiveness or causal superiority because allocation was quasi-experimental and both groups received credible chatbot-based lifestyle support. NExGEN’s automated, remotely delivered architecture offers scalability in principle, but population reach, implementation cost, staff burden, and effectiveness under routine campus conditions were not measured. Scalability should therefore be regarded as a testable implementation proposition rather than a demonstrated consequence of these results. ([JMIR][1])

The improvement in psychological well-being within the NExGEN group is broadly consistent with evidence that digitally delivered interventions can enhance mental health and well-being among university students. A systematic review of 89 college-student studies found that most digital programs produced beneficial or partly beneficial psychological changes, although risk of bias was often substantial and implementation outcomes were inconsistently reported (Lattie et al., 2019). Meta-analytic evidence has likewise indicated small-to-moderate benefits of internet interventions across student mental-health outcomes (Harrer et al., 2019). More directly, a guided online acceptance and commitment therapy program improved university-student well-being relative to a waiting-list comparator, supporting the plausibility of meaningful change through structured digital self-management (Räsänen et al., 2016). The present within-group result aligns with that general evidence but cannot identify which NExGEN component accounted for the change. NExGEN combined personalized onboarding, adaptive weekly actions, repeated check-ins, reminders, and multi-domain lifestyle content, so the estimated change cannot be assigned specifically to generative AI, personalization, or any single behavioral technique. The absence of a significant between-group contrast also differs from the preliminary Woebot trial, in which a conversational-agent intervention produced a greater reduction in depressive symptoms than an information-only control (Fitzpatrick et al., 2017). This divergence is readily explained by design and outcome differences. The Woebot study evaluated depressive symptoms over two weeks against a comparatively low-intensity information control, whereas the present study assessed positive psychological well-being over 12 weeks and compared two groups using the same chatbot environment, schedule, lifestyle domains, reminders, and technical support. Consistent with this explanation, Davies et al. (2014) reported that university digital interventions separated more clearly from inactive controls than from active comparators. The current confidence interval remains compatible with no incremental benefit and with a modest advantage for NExGEN, but it does not justify describing the result as a favorable trend. A definitive next trial should use individual randomization, retain the active structured-prompt arm, add a usual-care or assessment-only arm, and prespecify a minimally important between-group difference. Intermediate assessments and post-intervention follow-up would then clarify onset, trajectory, and durability. ([JMIR][2])

The body-appreciation finding similarly aligns with evidence that positive body image can be modified through structured psychological and behavioral content. Meta-analytic work found that stand-alone body-image interventions produced small average improvements and emphasized the need for larger, methodologically stronger trials (Alleva, Sheeran, et al., 2015). A systematic review focused specifically on positive body image found supportive evidence for approaches involving self-compassion, cognitive-behavioral strategies, intuitive-eating principles, and physical activity, while also identifying heterogeneity and limitations in study quality (Guest et al., 2019). NExGEN included body functionality, body respect, self-compassion, reduced appearance-based comparison, and safe lifestyle actions. One possible explanation for the significant within-group increase is that repeated exposure to these themes shifted attention from appearance evaluation toward appreciation of what the body can do. This mechanism is consistent with the Expand Your Horizon randomized trial, in which functionality-focused writing improved body appreciation and related outcomes compared with an active control (Alleva, Martijn, et al., 2015). It remains speculative in the present study because functionality appreciation, self-compassion, appearance comparison, and other candidate mediators were not tested as explanatory pathways. More importantly, the adjusted difference in body-appreciation change between NExGEN and the structured-prompt control was not statistically significant after Holm correction. The comparator also addressed body respect and received the same platform exposure and contact structure, so shared content may have reduced the contrast attributable to personalization. The study therefore extends positive-body-image intervention research to a Malaysian university context and to an AI-supported delivery format, but it does not show that personalization added benefit beyond structured chatbot guidance. A mechanism-focused trial should measure candidate mediators repeatedly and compare personalized and standardized versions of the same body-image modules, allowing the incremental contribution of personalization to be separated from the effect of receiving positive-body-image content at all. ([PLOS][3])

The two null between-group findings require interpretation as primary results rather than as secondary qualifications to the within-group changes. Neither null contrast demonstrates that the interventions were equivalent, because equivalence margins were not specified and the confidence intervals allowed a range of plausible incremental effects. Conversely, the significant changes observed within NExGEN cannot be used to infer superiority, particularly because change over time may reflect shared intervention components, assessment effects, secular influences, regression toward typical values, or uncontrolled differences between groups. The active control was methodologically demanding: participants used the same ChatGPT model and institutional environment, covered the same broad lifestyle domains, received the same orientation and reminder frequency, and completed weekly check-ins. This design strengthens the test of personalization itself, but it also makes separation more difficult because both conditions contain potentially beneficial ingredients. The findings therefore align more closely with the active-comparator pattern reported by Davies et al. (2014) than with studies comparing a digital intervention against minimal information or waiting-list controls (Fitzpatrick et al., 2017; Räsänen et al., 2016). The 12-week duration may also have been sufficient for changes common to both programs while remaining insufficient to reveal a small incremental effect of adaptive personalization. In addition, matched block allocation balanced prespecified characteristics but could not control all measured and unmeasured determinants of well-being, body appreciation, or digital engagement. These considerations do not convert the null findings into evidence for NExGEN; they identify why the incremental effect remains unresolved. A three-arm randomized trial should stratify by university, sex, and baseline outcome severity, maintain equal platform exposure across intervention arms, and extend follow-up beyond the active period. Such a design would distinguish change attributable to general chatbot-supported lifestyle guidance from change specifically associated with adaptive personalization and would provide a more direct test of the two research gaps identified for Malaysian university students. ([JMIR][1])

The theoretical implication is correspondingly narrower than a claim that personalization is inherently more effective. NExGEN operationalized personalization as a bundled system that transformed a 47-item assessment into a longitudinal context profile, generated individualized weekly actions, and adapted later content to reported progress and barriers. The significant within-group changes indicate that this package was compatible with improvement, but the null between-group contrasts do not isolate content specificity as the active ingredient. Common factors shared by both conditions, including structured self-monitoring, repeated prompts, consistent access, reminders, and exposure to supportive lifestyle content, may have contributed substantially. This interpretation is consistent with digital mental-health reviews that identify substantial heterogeneity in intervention components and call for studies that determine which elements account for benefit (Abd-Alrazaq et al., 2020; Lattie et al., 2019). Future evaluations should therefore move beyond whole-package comparisons. A factorial or micro-randomized design could vary onboarding depth, adaptive feedback, reminder timing, and personalized action selection independently. Such component-level testing would determine whether personalization changes outcomes directly, increases engagement, or works only for participants with particular baseline needs or barriers. ([JMIR][4])

Engagement and acceptability provide useful process context but do not alter the null comparative conclusions. Participants in the NExGEN group recorded a median of 68.00 platform logins (IQR, 59.00–74.00), compared with 58.50 logins (IQR, 52.00–70.50) in the control group. Mean acceptability scores were 3.92 (SD, 0.42) and 3.59 (SD, 0.41), respectively. These descriptive differences show numerically higher use and acceptability in NExGEN, but no hypothesis test was reported for either comparison; they should not be presented as statistically greater engagement or satisfaction. Nor can engagement be treated as a mediator of outcome change from these summaries. Engagement with digital interventions includes both observable behavior and subjective experience, and its relation to outcomes can be reciprocal: greater use may support change, while early perceived benefit may itself encourage continued use (Perski et al., 2017). A dedicated analysis of these usage patterns is [TO CONFIRM: planned as a separate publication]. That analysis should model weekly engagement trajectories rather than cumulative logins alone, distinguish exposure from active participation, and test whether engagement precedes subsequent outcome change. Any mediation analysis should account for baseline motivation, prior ChatGPT use, and time-varying outcome status, while retaining the exploratory status appropriate to nonrandomized mediator data. These steps would clarify whether personalization affects outcomes through engagement, whether acceptability and use merely mark participants already likely to improve, or whether engagement contributes little once shared intervention exposure is considered. ([OUP Academic][5])

Several limitations define the strength of the conclusions. First, the quasi-experimental allocation procedure leaves residual confounding possible despite matched blocks, covariate adjustment, and propensity-score sensitivity planning; causal attribution to NExGEN is therefore not warranted. Second, participants and the intervention coordinator could not be blinded, creating expectancy and differential-motivation effects. Blinding outcome assessors and the statistician reduced, but could not remove, this risk. Third, the active control was both a strength and a constraint. Matching the platform, duration, lifestyle domains, reminders, technical support, and safety procedures isolated the incremental value of personalization more rigorously than an inactive control, but likely reduced the detectable contrast. Fourth, outcomes were measured only at baseline and week 12. The design could not identify when change occurred, whether it persisted, or whether engagement preceded outcome improvement. Fifth, WHO-5 and BAS-2 scores were self-reported and may have been influenced by expectancy or socially desirable responding, although validated language versions and blinded assessment procedures strengthened measurement consistency. Sixth, incomplete week-12 data required inference under missing-data assumptions that cannot be verified fully. Finally, recruitment from three universities in one Malaysian district, together with requirements for device access and digital literacy, limits generalization to students in other regions, students with limited connectivity, and clinical populations excluded for safety reasons. Strengths included an intention-to-treat framework, multiplicity control for co-primary outcomes, an exposure-matched comparator, objective platform logs, fidelity monitoring, and blinded outcome assessment and analysis. Overall, NExGEN participation was associated with significant within-group improvements in psychological well-being and body appreciation, but the study did not demonstrate greater improvement than structured chatbot guidance. The engagement and acceptability summaries support continued evaluation, not a claim of comparative effectiveness. A longer, individually randomized trial with component-level testing is required before personalized NExGEN delivery can be recommended as superior to standardized AI-supported lifestyle guidance.

## Declarations

## Ethics Approval

REC/1954/2026

## Consent

Written informed consent was obtained from all participants.

## Data Availability

Data are available from the corresponding author on reasonable request

## Author Contributions

conceptualisation, methodology, writing original draft, KL: formal analysis.

## Use of Generative AI and AI-Assisted Technologies

During the preparation of this work, the authors used ChatGPT 5.4 for [AREAS OF THE MANUSCRIPT — e.g. drafting and language editing of the Introduction and Discussion]. All AI-generated content has been thoroughly reviewed, edited, and supplemented by the authors to ensure accuracy, originality, and alignment with the journal’s standards. The authors take full responsibility for all content presented in this submission.

## Data Availability

All data produced in the present study are available upon reasonable request to the authors

https://docs.google.com/spreadsheets/d/1K39KMobpfi6Isg74TWrqxwvcuTuOn37B/edit?usp=sharing&ouid=110563557379484039857&rtpof=true&sd=true

## References

1. Abd-Alrazaq, A. A., Rababeh, A., Alajlani, M., Bewick, B. M., & Househ, M. (2020). Effectiveness and safety of using chatbots to improve mental health: Systematic review and meta-analysis. Journal of Medical Internet Research, 22(7), e16021. 10.2196/16021

2. Alleva, J. M., Martijn, C., Van Breukelen, G. J. P., Jansen, A., & Karos, K. (2015a). Expand Your Horizon: A programme that improves body image and reduces self-objectification by training women to focus on body functionality. Body Image, 15, 81–89. 10.1016/j.bodyim.2015.07.001

3. Alleva, J. M., Sheeran, P., Webb, T. L., Martijn, C., & Miles, E. (2015b). A meta-analytic review of stand-alone interventions to improve body image. PLOS ONE, 10(9), e0139177. 10.1371/journal.pone.0139177

4. Auerbach, R. P., Mortier, P., Bruffaerts, R., Alonso, J., Benjet, C., Cuijpers, P., Demyttenaere, K., Ebert, D. D., Green, J. G., Hasking, P., Murray, E., Nock, M. K., Pinder-Amaker, S., Sampson, N. A., Stein, D. J., Vilagut, G., Zaslavsky, A. M., Kessler, R. C., & WHO WMH-ICS Collaborators. (2018). WHO World Mental Health Surveys International College Student Project: Prevalence and distribution of mental disorders. Journal of Abnormal Psychology, 127(7), 623–638. 10.1037/abn0000362

5. Braun, V., & Clarke, V. (2021). One size fits all? What counts as quality practice in (reflexive) thematic analysis? Qualitative Research in Psychology, 18(3), 328–352. 10.1080/14780887.2020.1769238

6. Conboy, L., Mingoia, J., Hutchinson, A. D., Reisinger, B. A. A., & Gleaves, D. H. (2024). Digital body image interventions for adult women: A meta-analytic review. Body Image, 51, 101776. 10.1016/j.bodyim.2024.101776

7. Davies, E. B., Morriss, R., & Glazebrook, C. (2014). Computer-delivered and web-based interventions to improve depression, anxiety, and psychological well-being of university students: A systematic review and meta-analysis. Journal of Medical Internet Research, 16(5), e130. 10.2196/jmir.3142

8. Des Jarlais, D. C., Lyles, C., Crepaz, N., & the TREND Group. (2004). Improving the reporting quality of nonrandomized evaluations of behavioral and public health interventions: The TREND statement. American Journal of Public Health, 94(3), 361–366. 10.2105/AJPH.94.3.361

9. Faul, F., Erdfelder, E., Buchner, A., & Lang, A.-G. (2009). Statistical power analyses using G*Power 3.1: Tests for correlation and regression analyses. Behavior Research Methods, 41(4), 1149–1160. 10.3758/BRM.41.4.1149

10. Fitzpatrick, K. K., Darcy, A., & Vierhile, M. (2017). Delivering cognitive behavior therapy to young adults with symptoms of depression and anxiety using a fully automated conversational agent (Woebot): A randomized controlled trial. JMIR Mental Health, 4(2), e19. 10.2196/mental.7785

11. Guest, E., Costa, B., Williamson, H., Meyrick, J., Halliwell, E., & Harcourt, D. (2019). The effectiveness of interventions aiming to promote positive body image in adults: A systematic review. Body Image, 30, 10–25. 10.1016/j.bodyim.2019.04.002

12. Harrer, M., Adam, S. H., Baumeister, H., Cuijpers, P., Karyotaki, E., Auerbach, R. P., Kessler, R. C., Bruffaerts, R., Berking, M., & Ebert, D. D. (2019). Internet interventions for mental health in university students: A systematic review and meta-analysis. International Journal of Methods in Psychiatric Research, 28(2), e1759. 10.1002/mpr.1759

13. Harris, P. A., Taylor, R., Thielke, R., Payne, J., Gonzalez, N., & Conde, J. G. (2009). Research electronic data capture (REDCap)—A metadata-driven methodology and workflow process for providing translational research informatics support. Journal of Biomedical Informatics, 42(2), 377–381. 10.1016/j.jbi.2008.08.010

14. Hashim, N., Hadi Munir, N. A. N., Ahmad Tarmizi, N. A., & Ahmad, N. F. I. (2022). Dissatisfaction about body image during social networking among university students. The Malaysian Journal of Nursing, 13(4), 12–18. 10.31674/mjn.2022.v13i04.003

15. Lattie, E. G., Adkins, E. C., Winquist, N., Stiles-Shields, C., Wafford, Q. E., & Graham, A. K. (2019). Digital mental health interventions for depression, anxiety, and enhancement of psychological well-being among college students: Systematic review. Journal of Medical Internet Research, 21(7), e12869. 10.2196/12869

16. Madrid-Cagigal, A., Kealy, C., Potts, C., Mulvenna, M. D., Byrne, M., Barry, M. M., & Donohoe, G. (2025). Digital mental health interventions for university students with mental health difficulties: A systematic review and meta-analysis. Early Intervention in Psychiatry, 19(3), e70017. 10.1111/eip.70017

17. Mahon, C., & Seekis, V. (2022). Systematic review of digital interventions for adolescent and young adult women’s body image. Frontiers in Global Women’s Health, 3, 832805. 10.3389/fgwh.2022.832805

18. Nyakhar, S., & Wang, H. (2025). Effectiveness of artificial intelligence chatbots on mental health & well-being in college students: A rapid systematic review. Frontiers in Psychiatry, 16, 1621768. 10.3389/fpsyt.2025.1621768

19. Perski, O., Blandford, A., West, R., & Michie, S. (2017). Conceptualising engagement with digital behaviour change interventions: A systematic review using principles from critical interpretive synthesis. Translational Behavioral Medicine, 7(2), 254–267. 10.1007/s13142-016-0453-1

20. Räsänen, P., Lappalainen, P., Muotka, J., Tolvanen, A., & Lappalainen, R. (2016). An online guided ACT intervention for enhancing the psychological wellbeing of university students: A randomized controlled clinical trial. Behaviour Research and Therapy, 78, 30–42. 10.1016/j.brat.2016.01.001

21. Suhaimi, A. F., Makki, S. M., Tan, K.-A., Silim, U. A., & Ibrahim, N. (2022). Translation and validation of the Malay version of the WHO-5 Well-Being Index: Reliability and validity evidence from a sample of type 2 diabetes mellitus patients. International Journal of Environmental Research and Public Health, 19(7), 4415. 10.3390/ijerph19074415

22. Swami, V., Mohd. Khatib, N. A., Toh, E., Zahari, H. S., Todd, J., & Barron, D. (2019). Factor structure and psychometric properties of a Bahasa Malaysia (Malay) translation of the Body Appreciation Scale-2 (BAS-2). Body Image, 28, 66–75. 10.1016/j.bodyim.2018.12.006

23. Tan, C.-S., Cheng, S.-M., Cong, C. W., Abu Bakar, A. R. B., Michael, E., Mohd Wazir, M. I. B., Mat Alim, M. B., Ahmad Tajudin, B. D. B., Mohamed Rosli, N. E.-Z. H. B., & Asmi, A. B. (2021). Validation and measurement invariance of the Body Appreciation Scale-2 between genders in a Malaysian sample. International Journal of Environmental Research and Public Health, 18(21), 11628. 10.3390/ijerph182111628

24. Topp, C. W., Østergaard, S. D., Søndergaard, S., & Bech, P. (2015). The WHO-5 Well-Being Index: A systematic review of the literature. Psychotherapy and Psychosomatics, 84(3), 167–176. 10.1159/000376585

25. Tylka, T. L., & Wood-Barcalow, N. L. (2015). The Body Appreciation Scale-2: Item refinement and psychometric evaluation. Body Image, 12, 53–67. 10.1016/j.bodyim.2014.09.006

26. White, I. R., Royston, P., & Wood, A. M. (2011). Multiple imputation using chained equations: Issues and guidance for practice. Statistics in Medicine, 30(4), 377–399. 10.1002/sim.4067

27. Wong, S. S., Wong, C. C., Ng, K. W., Bostanudin, M. F., & Tan, S. F. (2023). Depression, anxiety, and stress among university students in Selangor, Malaysia during COVID-19 pandemics and their associated factors. PLOS ONE, 18(1), e0280680. 10.1371/journal.pone.0280680

28. Zhong, W., Luo, J., & Zhang, H. (2024). The therapeutic effectiveness of artificial intelligence-based chatbots in alleviation of depressive and anxiety symptoms in short-course treatments: A systematic review and meta-analysis. Journal of Affective Disorders, 356, 459–469. 10.1016/j.jad.2024.04.057

